# Partner Military Deployment During Wartime Is Associated with Maternal Depression and Impaired Bonding: A Matched-Control Study from the Israel-Hamas War

**DOI:** 10.1101/2025.01.20.25320861

**Authors:** Hadas Allouche-Kam, Sabrina J Chan, Isha H Arora, Christina T Pham, Inbal Reuveni, Eyal Sheiner, Sharon Dekel

## Abstract

Purpose. The pregnancy and postpartum periods represent a time of heightened psychological vulnerability with implications for the offspring. Knowledge of the mental health of perinatal women exposed to armed conflict when their partner is in military deployment is scarce. Methods. This matched-control, survey-based study included a sample of 429 women recruited during the first months of the Israel-Hamas War who were pregnant or within six months postpartum. Women who had a partner in military deployment were matched primarily on demographics, prior mental health, and trauma exposure to women whose partner was no longer deployed. Results. We found that nearly 44% of pregnant women with a partner deployed endorsed probable depression. This group was more than twice as likely to endorse probable depression than matched pregnant controls. Likewise, postpartum women with a partner deployed reported significantly more maternal-infant attachment problems than the matched postpartum group of partners not deployed. Importantly, analysis showed that partner’s active deployment was related to maternal depression and attachment problems via reduced perceived social support. Conclusions. Partner military deployment during conditions of war can serve as a major psychological stressor for pregnant and postpartum women. It can heighten psychiatric morbidity and interfere with attachment to the infant in part by diminished social support. Implementation of community-based services for the peripartum population is crucial during times of war and other large-scale traumas.

**Article Highlights:** Partner military deployment increases risk for antepartum depression and maternal-infant bonding problems.

Reduced social support explains these maternal outcomes.

Clinical attention to the wellbeing of the peripartum population is warranted during times of collective trauma.

## Introduction

On October 7, 2023, the Israel-Hamas conflict escalated when the Hamas organization infiltrated southern Israel through coordinated land, sea, and air attacks. Israeli civilians residing in communities near the border, Nova festival participants, and soldiers were murdered, raped, and taken hostage, without distinction between children and elderly or Jewish and non-Jewish victims (Secretary-General of the United Nations 2024). This event, classified as a war crime by the UN Human Rights Council (Office of the United Nations High Commissioner for Human Rights 2024), resulted in over 1,250 fatalities and 251 hostages (U.S. Department of Defense; Levany et al. 2023; Secretary-General of the United Nations 2024). In response, Israel declared war, and the Israeli Defense Forces (IDF) invaded Gaza, striking multiple Hamas targets. There was a rapid and massive deployment of reserve duty by the IDF, with many soldiers serving on active duty for months.

Mental health effects of man-made trauma, such as war and terrorism, on individuals directly exposed, those indirectly exposed, and the community at large have been extensively studied (Silver 2002; Yehuda and Hyman 2005; Perlman et al. 2011; Morina et al. 2018; Castro-Vale et al. 2019; Brown 2020; Hoppen et al. 2021). Man-made trauma shatters people’s core cognitive schemas that the world is a predictable and safe place (Chrisman and Dougherty 2014). The amplitude of studies report ensuing psychiatric conditions among direct and indirect trauma-exposed individuals (Punamaki et al. 2018; Druetz et al. 2020; Klapper-Goldstein et al. 2024; Krupelnytska and Morozova-Larina 2025) — most notably post-traumatic stress disorder (PTSD) and depression, which may critically and persistently undermine the person’s physical health (VanItallie 2002; Glaesmer et al. 2011; Nemeroff and Goldschmidt-Clermont 2012; Roberts et al. 2015; Rosenbaum et al. 2015; Perkins et al. 2021). In the general community, an estimated 1 in 4 individuals residing in a war-conflict zone will be affected by PTSD (Ahmed et al. 2024) with potential intergenerational effects on the offspring, underscoring the magnitude of psychiatric morbidity in exposed communities (Devakumar et al. 2014; Weinstein et al. 2018; Yehuda and Lehrner 2018; Castro-Vale et al. 2019; Brown 2020; Hoppen et al. 2021). Not surprisingly, a national survey revealed elevated mental health complications among the Israeli community following the October 7th, 2023, attack (Levi-Belz et al. 2024; Hasson-Ohayon and Horesh 2024).

There is a critical gap in knowledge concerning the effects of war exposure on the perinatal population. Pregnancy and the postpartum period have been repeatedly documented as times of potential psychological vulnerability (Louise M. Howard 2020; Wisner et al. 2024). Perinatal depression is the leading pregnancy complication, affecting 1 out of 7 women residing in non-war-conflict zones (Gavin et al. 2005; Curry et al. 2019). It increases risk for maternal (Ackerman‐Banks et al. 2023; Meaidi 2024), fetus (Alder et al. 2007; Grote et al. 2010), and infant health complications (Jarde et al. 2016; Smith et al. 2020; Rogers et al. 2020; Simonovich et al. 2021). In its extreme form, it can result in maternal death and infanticide (Jones et al. 2014). Although maternal stress has been linked to poor childbirth outcomes, studies on the adversity of war and terrorism on perinatal mental health are largely lacking (Yehuda et al. 2005; Engel et al. 2005; Klapper-Goldstein et al. 2024; Krupelnytska and Morozova-Larina 2025).

An important factor to consider in understanding the unique impact of war on the perinatal population is partner military deployment. Partners often serve as the primary source of emotional reassurance and practical aid (Houts et al. 2008). Lack of partner support increases risk for perinatal depression and maternal-infant attachment problems (Bilszta et al. 2008; Biaggi et al. 2016). Although the literature is in its infancy, existing studies on partner deployment reveal substantial depression in women residing in non-war zones (Razurel et al. 2013; Tarney et al. 2015; Godier-McBard et al. 2019; Pretorius et al. 2024).

A large body of research underscores the adaptive role of social support in coping following a traumatic event (Wickramaratne et al. 2022; Calhoun et al. 2022). Post-trauma recovery has been understood within an interpersonal framework, whereby relational partners facilitate coping through the provision of social support (Calhoun et al. 2022). Partner deployment may act as a stressor that heightens feelings of helplessness and loneliness while depleting psychological resources needed to cope with trauma exposure during the demanding perinatal period (Robrecht et al. 2008; Klaman and Turner 2016; Godier-McBard et al. 2019; Pretorius et al. 2024). To our knowledge, no study has examined the impact of partners’ deployment on mental health and maternal-infant attachment among pregnant and postpartum women who have been exposed, both directly and indirectly, to war-related stressors.

We studied a sample of 165 pregnant and 264 postpartum women in the first months following the events of October 7th and the outbreak of the war, including some who had partners actively deployed. We asked whether partner deployment is associated with maternal mental health and maternal-infant attachment, and whether social support mediates the relationship between partner deployment and maternal outcomes.

## Materials and Methods

The study commenced on January 19th, 2024, during the first months of the Israel-Hamas (“Iron Swords”) War to understand the impact of terrorism and war on maternal health (n = 1,085). In the months leading up to study recruitment (i.e., October 2023-January 2024), over 10,600 rockets and mortar rounds were launched toward Israel and during the recruitment, there were 425 missile alerts across the country (The Meir Amit Intelligence and Terrorism Information Center 2024; Israel’s National Emergency Portal 2024) and 10 terrorist attacks (The Meir Amit Intelligence and Terrorism Information Center 2024).

Women residing in Israel during the war who were pregnant or gave birth within the past six months were recruited via social media and professional organizations. They were asked to complete a survey about mental health, maternal-infant attachment, trauma exposure including partner deployment, and obstetrics and childbirth-related factors. The study was approved by Mass General Brigham Human Research Committee. Participants provided implied consent by providing responses in study questionnaires.

A total of 466 (42.9%) of participants reported partner deployed since October 7th, 2023, either currently or no longer. 7.9% were missing information on important study outcomes, resulting in a sample of 429 participants. This includes 165 (38.5%) pregnant women and 264 (61.5%) postpartum women. 99.3% were Jewish. Information regarding the sample characteristics is provided in Table 1.

**Table 1.** Socio-demographics, trauma exposure, and obstetrical information for pregnant and postpartum women by partner deployment status.

|  | Pregnant Women (N = 165) |  | Postpartum Women (N = 264) |  |
| --- | --- | --- | --- | --- |
|  | Active Partner<br>Deployment | Non-Active<br>Partner<br>Deployment | Active Partner<br>Deployment | Non-Active<br>Partner<br>Deployment |
|  | n (%) / M (SD) | n (%) / M (SD) | n (%) / M (SD) | n (%) / M (SD) |
| **Demographic Factors** |  |  |  |  |
| Maternal Age | 31.86 ± 4.06 | 31.31 ± 4.43 | 31.79 ± 4.55 | 32.22 ± 4.05 |
| Mental Illness before October 7th | 9 (8.41) | 2 (3.45) | 10 (6.99) | 14 (11.57) |
| Marital Status |  |  |  |  |
| Married or domestic partnership | 106 (99.07) | 57 (98.28) | 142 (99.3) | 121(100.00) |
| Single, in a relationship | 1 (0.93) | 1 (1.72) | 1 (0.70) | 0 |
| Employment |  |  |  |  |
| Employed | 75 (70.09) | 45 (78.95) | 97 (68.31) | 91 (75.83) |
| Home-employed | 10 (9.35) | 4 (7.02) | 10 (7.04) | 7 (5.83) |
| Self-employed | 10 (9.35) | 4 (7.02) | 15 (10.56) | 13 (10.83) |
| Student | 11 (10.28) | 4 (7.02) | 12 (8.45) | 5 (4.17) |
| Unemployed | 1 (0.93) | 0 | 8 (5.63) | 4 (3.33) |
| Education |  |  |  |  |
| High School or equivalent | 5 (4.67) | 3 (5.17) | 5 (3.50) | 2 (1.65) |
| Some college | 5 (4.67) | 1 (1.72) | 7 (4.90) | 8 (6.61) |
| Bachelor’s Degree | 57 (53.27) | 36 (62.07) | 78 (54.55) | 56 (46.28) |
| Master’s Degree | 34 (31.76) | 15 (25.86) | 43 (30.07) | 46 (38.02) |
| Doctoral or Medical Degree | 6 (5.61) | 3 (5.17) | 10 (6.99) | 9 (7.44) |
| Household Income (Monthly) |  |  |  |  |
| Up to 7106 NIS | 7 (6.54) | 4 (6.90) | 7 (4.90) | 8 (6.61) |
| Up to 12862 NIS | 20 (18.69) | 11 (18.97) | 36 (25.17) | 29 (23.97) |
| Up to 22847 NIS | 46 (42.99) | 26 (44.83) | 57 (39.86) | 33 (27.27) |
| Over 22847 NIS | 34 (31.78) | 17 (29.31) | 43 (30.07) | 51 (42.25) |
| **Trauma Exposure Factors** |  |  |  |  |
| Home Zones |  |  |  |  |
| Red | 9 (8.41) | 5 (8.62) | 15 (10.49) | 9 (7.44) |
| Being Displaced | 2 (22.22) | 1 (20.00) | 4 (26.67) | 1 (11.11) |
| Orange | 27 (25.23) | 13 (22.41) | 35 (24.48) | 32 (26.45) |
| Yellow | 30 (28.04) | 10 (17.24) | 35 (24.48) | 30 (24.79) |
| Green | 41 (38.32) | 30 (51.72) | 58 (40.56) | 50 (41.32) |
| Watch uncensored videos (past week) | 7 (6.54) | 0 | 12 (8.39) | 12 (9.92) |
| Know civilians murdered | 42 (39.25) | 20 (34.48) | 61 (42.66) | 39 (32.23) |
| Know civilians and/or soldiers taken<br>hostage | 16 (14.95) | 7 (12.07) | 25 (17.48) | 21 (17.36) |
| **Obstetrics Factors** |  |  |  |  |
| Primiparity | 25 (23.36) | 17 (29.31) | 36 (25.17) | 43 (35.54) |
| Gestational age |  |  |  |  |
| 1<sup>st</sup> Trimester | 23 (21.49) | 5 (8.62) | - | - |
| 2<sup>nd</sup> Trimester | 39 (36.45) | 25 (43.10) | - | - |
| 3<sup>rd</sup> Trimester | 45 (42.06) | 28 (48.28) | - | - |
| Birth after October 7th | - | - | 86 (60.14) | 65 (53.72) |
| Mode of Delivery |  |  |  |  |
| Vaginal delivery | - | - | 106 (79.70) | 85 (71.43) |
| Assisted vaginal delivery | - | - | 11 (8.27) | 8 (6.72) |
| Scheduled caesarean delivery | - | - | 6 (4.51) | 11 (9.24) |
| Emergent caesarean delivery | - | - | 10 (7.52) | 15 (12.61) |
| NICU admittance | - | - | 9 (6.82) | 8 (6.72) |

Note. NIS, New Israeli Shekel. All exposure factors related to the October 7th events and the war. Home Zone refers to geographic zones as defined by the Israeli Defense Forces (IDF) Home Front Command to indicate the available time to reach shelter during missile attacks: Red ( $\leq 15$ seconds), Orange (30 seconds), Yellow (45 seconds), and Green (60-90 seconds). Watched uncensored videos in the past week; Values pertain to high frequency of exposure (score 8 or above on a 1-10 scale regarding amount (hours) of exposure daily). Gestational age was reported according to trimesters (weeks) (0-13, first trimester; 14-27, second trimester; 28-42, third trimester). NICU = Neonatal Intensive Care Unit.

Geographic exposure zones were classified based on IDF Home Front Commands: Red, Orange, Yellow and Green, defined by time available to reach shelter for threat of rocket and missile attacks from the time of the siren (i.e., immediate to 15 seconds, i.e., highest level of proximity to combat zone, 30, 45, and 60-90 seconds). The majority of the sample resided during October 7th in the Green zone (41.7%), 24.9% and 24.5% in the Orange and Yellow zones, respectively, and 8.9% in the Red zone with 21.05% displaced. 74.1% of participants knew people who were murdered, killed, injured, and/or taken hostage as a result of October 7th and the war.

## Measures

Depression symptoms were assessed using the Edinburgh Postnatal Depression Scale (EPDS), a 10-item questionnaire assessing depression symptoms over the past seven days (Cox et al. 1987). Higher scores indicate a greater level of symptoms. The EPDS is the recommended screening tool in perinatal settings, as determined by American College of Obstetricians and Gynecologists (ACOG) (American College of Obstetricians and Gynecologists 2023) and the US Preventive Services Task Force (Curry et al. 2019). We used the cutoff score of 14 to indicate probable depression (Cox et al. 1987). Reliability was high (α = 0.86).

Posttraumatic stress disorder (PTSD) symptoms were measured with the well-validated PTSD Checklist for DSM-5 (PCL-5)(Weathers et al. 2013). This self-report tool is designed to assess the severity of post-traumatic stress disorder (PTSD) symptoms in the past month in regard to a specified trauma (here, October 7th events) and recommended by the Veterans of Affairs (VA) for provisional PTSD (Weathers 2013; Bovin et al. 2016). It lists the 20 DSM-5 PTSD symptoms. Higher scores indicate higher symptom levels. We used the cutoff score of 33 to determine probable PTSD (Bovin et al. 2016) (α = 0.91).

Maternal-fetal attachment in pregnancy was measured using the Maternal-Fetal Attachment Scale (MFAS) (Cranley 1981). This is a 24-item questionnaire designed to evaluate the emotional connection between a mother and her fetus. Higher scores indicate greater maternal attachment. It shows strong psychometric properties (Lindgren 2001; Wittkowski et al. 2020) (α = 0.88).

Maternal-infant attachment during the postpartum period was assessed using the Maternal Attachment Inventory (MAI) (Müller 1994). This 26-item self-report scale evaluates a mother’s perceptions and emotions regarding her infant during the first year postpartum. It accords with observational data of bond behavior (Wittkowski et al. 2020). Higher scores indicate attachment problems (α = 0.94).

The Multidimensional Scale of Perceived Social Support (MSPSS) is a widely used 12-item questionnaire to assess perceived social support with higher scores indicating greater support. The scale demonstrates strong reliability and validity across trauma-exposed and peripartum populations (Dambi et al. 2018; Bedaso et al. 2021) (α = 0.93).

Socio-demographics, prior mental health problems, and obstetric information (e.g. gestational week, giving birth before or after the war, delivery mode, neonatal intensive care unit (NICU) admission) were measured by single items. Trauma exposure included information about participants’ place of residence on October 7th, defined by Home Front Command zones, whether participants knew people murdered, taken hostage, or injured, and degree of exposure to October 7th events/war via social media (i.e., exposure to uncensored movies).

## Data Analysis

Demographic data for maternal age and Home Front Commands exposure zone were noted to be missing at random, and multiple imputations using the *mice* R package (Buuren and Groothuis-Oudshoorn 2011) were performed. We conducted a full propensity score procedure using *MatchIt* R package (Ho et al. 2011) for pregnant and postpartum participants, separately, and matched participants whose partner was in active military deployment (study group) and participants whose partner was no longer in active deployment (control group). Propensity scores were estimated using logistic regression, modeling the probability of being in the study group as a function of selected baseline characteristics. Full matching was applied, which retained all participants by constructing matched sets where each set contained at least one individual from the study group and at least one from the control group. Participants were assigned weights based on the structure of matched sets, and all statistical analyses incorporated these weights. Groups were matched on demographics (i.e., maternal age, education, income), primiparity, prior mental health, exposure to October 7th and the war (i.e., residing in the Red zone, knowing civilians murdered and/or civilians/soldiers taken hostage, and watching related uncensored videos). Pregnant women were also matched on gestational week period and postpartum women on whether they gave birth before or after October 7th.

Following the matching, we compared groups on depression and PTSD, maternal-infant attachment, and social support. Kolmogorov-Smirnov and Shapiro-Wilk tests revealed that all factors in consideration were significantly positively skewed (p < 0.001). Analyses were conducted separately for pregnant and postpartum women. To estimate differences between women with a partner currently in active deployment or not, Mann-Whitney U tests were performed for quantitative measures and logistic regressions for estimating differences in categorical measures. Confidence Intervals (CIs) for the tests were based on bootstrapping of 1000 resampling cycles. All analyses were conducted in R (R Core Team 2023).

To examine whether social support mediated the effects of study group (women with versus without a partner who is deployed) on maternal outcomes, we conducted a mediation model using observed measures within a Structural Equation Modeling (SEM) framework, estimating a saturated model in *lavaan* R package(Rosseel 2012). Significance was estimated by bias-corrected bootstrap analysis with 2000 sampling cycles. Missing data were handled with the Full Information Maximum Likelihood (FIML) procedure. The group (0 = partner in non-active deployment, 1 = partner in active deployment) served as the predictor, social support as the mediator, and peripartum depression and attachment problems as the outcomes in separate analyses.

## Results

The rate of clinically significant depression symptoms (≥ 14, EPDS), indicative of probable depression, in pregnant women with partners in active deployment was 43.96% in comparison to 25.49% in the matched group of partners not deployed and represented a two-fold increase (χ^2^ = 4.01, Odds Ratio, OR = 2.29, 95% CI = 1.10 – 5.00, p = 0.05). No differences were found in depression rates among postpartum women with and without a partner deployed and in probable PTSD rates for pregnant and postpartum women by deployment status (Table 2). The depression rate in the pregnant active partner deployed group was also higher than in postpartum women with a partner deployed (χ^2^ = 8.51, OR = 2.46, 95% CI = 1.38 – 4.44, p = 0.00).

**Table 2.** Maternal depression, mother-infant bonding, and social support by partner deployment status.

|  | Pregnant Women (N = 165) |  |  |  |  | Postpartum Women (N = 264) |  |  |  |  | OR / Effect Size<br>(95% CI) |
| --- | --- | --- | --- | --- | --- | --- | --- | --- | --- | --- | --- |
| | Active Partner<br>Deployment | | Non-Active Partner<br>Deployment | | $\chi^2$ / Z | Active Partner<br>Deployment | | Non-Active Partner<br>Deployment | | $\chi^2$ / Z | |
|  | %/M | n/SD | %/M | n/SD |  | %/M | n/SD | %/M | n/SD |  |  |
| Probable<br>Depression | 43.96 | 40 | 25.49 | 13 | 4.01* | 23.89 | 27 | 23.08 | 24 | 0 | 2.29<br>(1.10 – 5.00)<sup>1</sup> |
|  |  |  |  |  |  |  |  |  |  |  | 1.05<br>(0.56 – 1.97)<sup>2</sup> |
| Probable<br>PTSD | 20.24 | 17 | 19.15 | 9 | 1.8e<sup>-30</sup> | 21 | 21 | 20 | 18 | 0 | 1.07<br>(0.44 – 2.73)<sup>1</sup> |
|  |  |  |  |  |  |  |  |  |  |  | 1.06<br>(0.52 – 2.17)<sup>2</sup> |
| Social<br>Support | 45.76 | 9.60 | 50.84 | 6.11 | 3.16** | 47.53 | 9.65 | 52.12 | 6.98 | 3.07** | 0.25 (0.10 – 0.38)<sup>1</sup> |
|  |  |  |  |  |  |  |  |  |  |  | 0.19 (0.07 – 0.30)<sup>2</sup> |
| Social<br>Support<br>(Significant<br>Other) | 15.73 | 3.36 | 17.86 | 1.93 | 3.97*** | 16.51 | 3.29 | 18.30 | 2.37 | 3.90*** | 0.31 (0.16 – 0.43)<sup>1</sup> |
|  |  |  |  |  |  |  |  |  |  |  | 0.24 (0.12 – 0.35)<sup>2</sup> |
|  | 15.16 | 3.96 | 16.82 | 2.50 | 2.25* | 15.48 | 4.04 | 17.03 | 3.36 | 2.62** | 0.17 (0.02 – 0.30)<sup>1</sup> |
| Social Support (Family) |  |  |  |  |  |  |  |  |  |  | 0.16 (0.04 – 0.28)<sup>2</sup> |
| Social Support (Friends) | 14.86 | 3.86 | 16.16 | 3.13 | 2.02* | 15.54 | 3.59 | 16.79 | 3.13 | 2.22* | 0.16 (0.02 – 0.30)<sup>1</sup> |
|  |  |  |  |  |  |  |  |  |  |  | 0.14 (0.02 – 0.26)<sup>2</sup> |
| Maternal Attachment | 61.65 | 14.00 | 63.02 | 15.12 | 0.61 | 32.44 | 7.28 | 30.27 | 5.50 | 2.36* | 0.05 (0.00 – 0.21)<sup>1</sup> |
|  |  |  |  |  |  |  |  |  |  |  | 0.15 (0.03 – 0.26)<sup>2</sup> |

Note. Probable depression was defined based on a score of 14 or higher on the Edinburgh Postnatal Depression Scale (EPDS $\geq 14$ ). Probable posttraumatic stress disorder (PTSD) regarding October 7th was defined based on a score of 33 or higher on the Posttraumatic Stress Disorder Checklist for DSM-5 (PCL-5 $\geq 33$ ). Maternal attachment refers to bonding with the fetus (higher scores better bonding) and bonding with the infant (higher scores worse attachment). M = Mean. SD = Standard deviation. OR = odd ratios, CI = confidence interval.
<sup>1</sup> = Active partner deployment v/s non-active partner deployment for pregnant women
<sup>2</sup> = Active partner deployment v/s non-active partner deployment for postpartum women
\*\*\*p<0.001, \*\*p<0.01, \*p<0.05

There were differences in maternal-infant attachment related to partner deployment. The active deployment group endorsed more attachment problems than controls (Z = 2.36, r = 0.15, CI = 0.03 – 0.26, p = 0.02) (Table 2), although there were no differences between the groups in delivery mode (χ^2^ = 4.83, p = 0.19) and NICU admission (χ^2^ = 7.5e^-30^, p = 1) (Table 1). No group differences were noted in maternal-fetal attachment (Table 2).

Both pregnant and postpartum women with a partner in active deployment reported significantly lower levels of social support than matched controls (Pregnant group: *Z* = 3.16, r = 0.25, CI = 0.1 – 0.38, p = 0.00; Postpartum group: *Z* = 3.07, r = 0.19, CI = 0.07 – 0.30, p = 0.00) (Table 2). For pregnant women, social support significantly mediated the path between study group and maternal depression (Figure 1). Partner in active deployment was related to lower social support (β = −0.30, p < 0.001), which in turn was associated with higher depression (β = 0.39, p <.001) (Figure 1). For postpartum women, social support mediated the path between study group and maternal-infant attachment, although the results were near significance. Partner in active deployment was related to lower social support (β = −0.26, p < 0.001), which in turn was associated with more attachment problems (β = −0.16, p < 0.1) (Figure 2).

**Figure 1.**
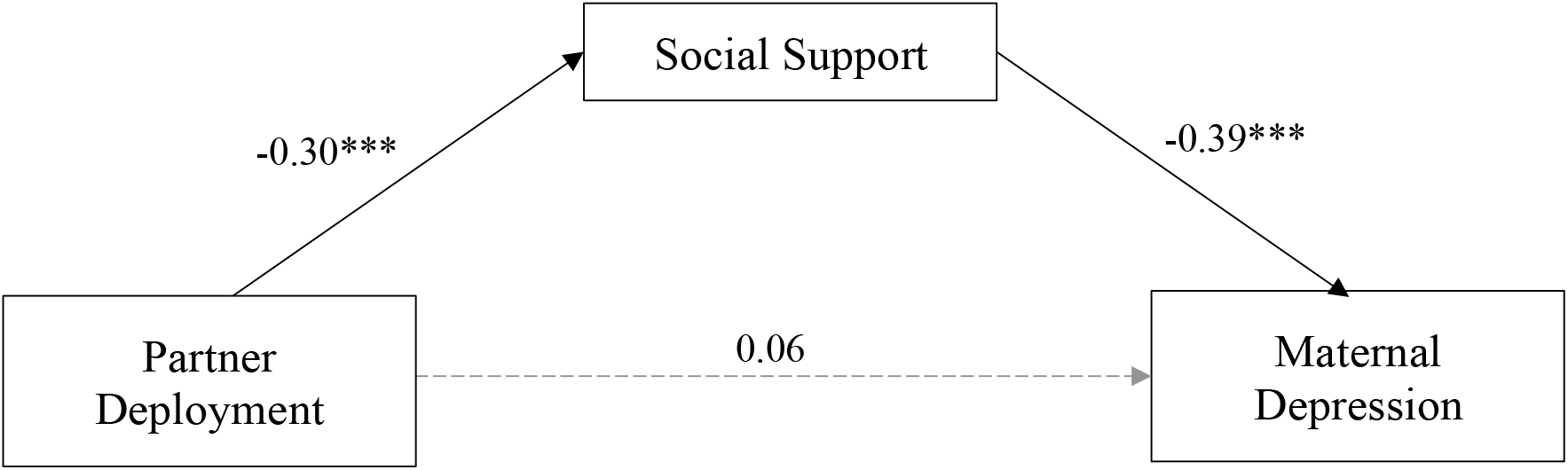
Associations between partner deployment, social support and antepartum depression. Mediation analysis linking study group (partner actively deployed v/s not) to symptoms of depression during pregnancy via perceived social support. Solid lines represent significant paths; Gray dashed lines represent non-significant paths. Values are standardized scores.

**Figure 2.**
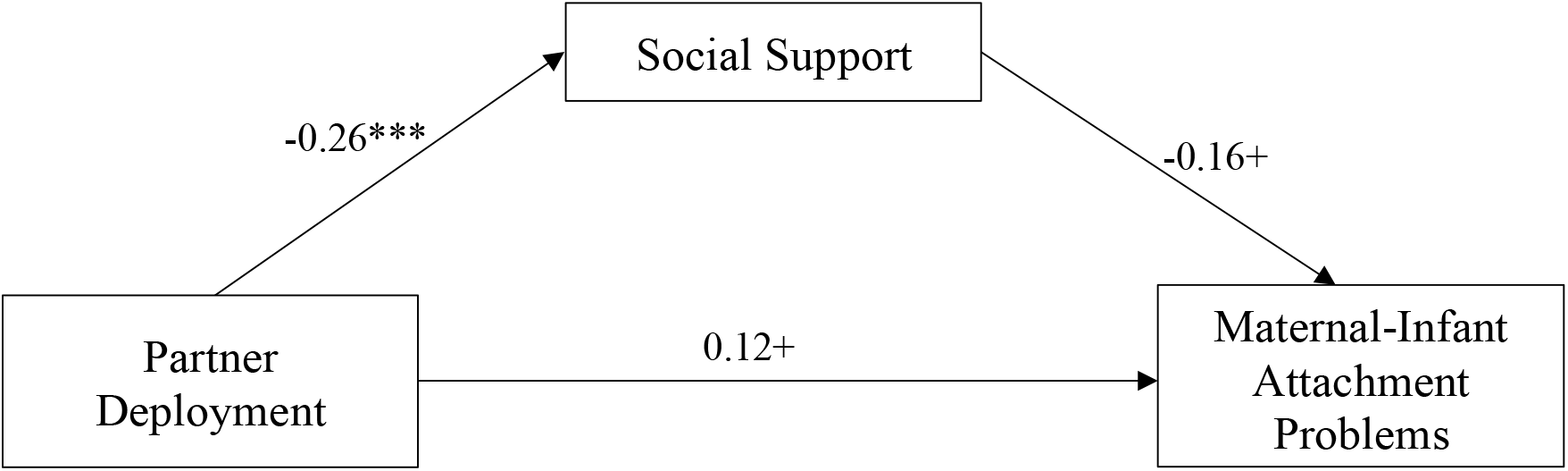
Associations between partner deployment, social support and mother-infant attachment problems. Mediation analysis linking study group (partner actively deployed v/s not) to maternal-infant attachment during the postpartum period via perceived social support. Solid lines represent significant or near significance paths. Values are standardized scores. ^***^p<0.001, ^+^<0.10

## Discussion and Conclusions

This study examined the psychological adjustment of pregnant and postpartum women exposed to war-related stressors during a time that their partner was in military deployment. Among pregnant women who were similar on demographic background, prior mental health, war-related exposure, and gestational week, those with a partner actively deployed were twice as likely to endorse probable depression compared to those with a partner no longer in active deployment. Similarly, postpartum women with a deployed partner reported greater difficulties bonding with their infant compared to those whose partner was not actively deployed, despite being comparable on other factors. As importantly, maternal adversity associated with having a partner deployed was mediated via reduced social support, underscoring the clinically significant impact of partner absence on maternal psychological health during times of collective trauma.

The findings reveal considerable psychological morbidity among perinatal women during wartime, with elevated rates of both depressive and posttraumatic symptoms. Under these conditions, partner deployment was associated with a markedly increased risk for probable depression among pregnant women, with prevalence approaching 44%. This accords with findings of elevated antepartum depression among American women living outside active war zones during their partner’s deployment (Haas et al. 2005; Smith et al. 2010; Levine et al. 2015; Tarney et al. 2015; Pretorius et al. 2024). A body of studies demonstrate the adverse impact of antepartum depression on maternal, fetus, and infant health (Bauer et al. 2016; Jarde et al. 2016; Smith et al. 2020; Simonovich et al. 2021; Ackerman‐Banks et al. 2023; Meaidi 2024). Depression increases risk for preterm labor, low birth weight and decreased head circumference (Engel et al. 2005; Ohlsson Arne 2011). It is linked with infant neurodevelopmental risks (Field 2011; Rogers et al. 2020; Fan et al. 2024; Miller et al. 2024) potentially via fetal programming, in which maternal distress contributes to the maternal-fetal biological milieu and, in turn, increases offspring’s susceptibility to disease states (Yehuda et al. 2005; Tegethoff et al. 2011; Yehuda and Lehrner 2018). Depression in pregnancy strongly associates with postpartum symptoms and depression in subsequent pregnancies (Milgrom et al. 2008; O’Hara and McCabe 2013).

To our knowledge, this study is the first to examine how partner deployment may undermine the formation of maternal-infant attachment. This bond is derived from daily, positive mother-infant interactions that are instrumental to the child’s emotional and social development (Moehler et al. 2006; O’Dea et al. 2023). While various mechanisms have been put forth for explaining the transmission of trauma across generations (Schore 2001; Yehuda et al. 2005; Lev-Wiesel 2007; Yehuda and Lehrner 2018; Bhattacharya et al. 2019), maternal-infant attachment may mediate the effect of maternal traumatic distress on the offspring’s welfare (Schore 2001; Benoit et al. 2010; Enlow et al. 2014; Ahlfs-Dunn et al. 2022). Stress and trauma may undermine maternal emotional regulation and maternal sensitive care (Enlow et al. 2014; Kumar et al. 2020; Dekel et al. 2020). This study further suggests that partner deployment may add to the impact of maternal trauma exposure on maternal-infant bonding.

The findings underscore the critical role of social support for maternal wellbeing during times of war. Partner deployment may substantially diminish women’s overall perceived social support, which in turn can adversely affect maternal mental health and attachment with the infant. A body of research has documented the positive role of social support in post-trauma psychological adaptation (Holt-Lunstad and Uchino 2015; Gariépy et al. 2016; Wickramaratne et al. 2022; Calhoun et al. 2022). In particular, emotional and instrumental support from partners has been identified as critical for maternal wellbeing in non-conflict contexts (Bilszta et al. 2008; Leahy‐Warren et al. 2012; Tanner Stapleton et al. 2012; Pilkington et al. 2015). These findings highlight the need for a more nuanced understanding of the pathways through which partner deployment during a vulnerable period influences maternal mental health, including direct effects via reduction of specific types of social support. Scalable interventions, such as telehealth-based mental health support, online maternal support groups, virtual partner involvement and community-based services, may provide alternative sources of support and help mitigate the adverse effects of partner absence (Kees and Rosenblum 2015; Nolan et al. 2019; Seagle et al. 2021; Hendrikx and Murphy 2021). Strengths of this study include the assessment of both pregnant and postpartum women exposed to twofold levels of adversity: from partner military deployment and from residing in war zones themselves. We also employed a match-controlled study design to understand the unique contribution of partner deployment to maternal outcomes. Shortcomings include the reliance on an internet-based, convenience sample. This approach enabled rapid recruitment of women under conditions of armed conflict, a population that would otherwise be difficult to enroll. Although the sample largely represents the Jewish Israeli population on demographics, there could be a selection bias, and the findings may reflect cultural-specific factors. We did not include clinical assessments; the self-reported mental health instruments accord strongly with diagnostic tools; however, there could be reporting bias. The use of a cross-sectional design precludes causal interpretations. Future longitudinal studies of perinatal women and their partners in various war zones are warranted to fully elucidate the impact of partner deployment on maternal mental health and their temporal relationships, as well as risk and protective factors.

In summary, among women with deployed partners, we find higher levels of probable depression during pregnancy, as well as lower maternal-infant attachment in the postpartum. Partner deployment adds a significant amount of stress by reducing social support and undermining maternal mental health during times of heightened stress, such as living under conditions of war. Because maternal wellbeing is a bedrock of a healthy society (Dekel et al. 2024), there is a critical need to ensure that resources for social support are in place and available. On a community level, attention to the welfare of perinatal women exposed to large-scale traumas is warranted.

## Data Availability

The datasets generated and/or analysed during the current study are not publicly available due to anticipated ongoing collection of data for this study but are available from the corresponding author on reasonable request.

## Author Contribution

HAK contributed to project management activities and wrote parts of the manuscript.

SC created the online survey.

IHA conducted the statistical analysis and wrote the statistical approach and results.

CP contributed to the manuscript writing and editing.

IR and ES served as study consultants.

SD is the principal investigator of the larger project. She designed the conceptual study paradigm, collected the data, supervised the manuscript preparation, and wrote the manuscript.

All authors read and approved the final manuscript.

## Data availability

The datasets generated and/or analyzed during the current study are available from the corresponding author on reasonable request.

